# Systemic administration of the antisense oligonucleotide NS-089/NCNP-02 for skipping of exon 44 in patients with Duchenne muscular dystrophy: study protocol for a phase I/II clinical trial

**DOI:** 10.1101/2023.02.06.23285500

**Authors:** Takami Ishizuka, Hirofumi Komaki, Yasuko Asahina, Harumasa Nakamura, Norio Motohashi, Eri Takeshita, Yuko Shimizu-Motohashi, Akihiko Ishiyama, Chihiro Yonee, Shinsuke Maruyama, Eisuke Hida, Yoshitsugu Aoki

## Abstract

**Aim:** The purpose of this study is to evaluate the safety and pharmacokinetics of the novel morpholino oligomer NS-089/NCNP-02 which can induce exon 44 skipping, in patients with DMD. Additionally, we aimed to identify markers predictive of therapeutic efficacy and determine the optimal dosing for future studies.

**Methods:** This is an open-label, dose-escalation, two-center phase I/II trial in ambulant patients with DMD, presence of an out-of-frame deletion, and a mutation amenable to exon 44 skipping. Part 1 is a stepwise dose-finding stage (4 weeks) during which NS-089/NCNP-02 will be administered intravenously at four dose levels once weekly (1.62, 10, 40, and 80 mg/kg); Part 2 is a 24-week evaluation period based on the dosages determined during Part 1. The primary (safety) endpoints are the results of physical examinations, vital signs, 12-lead electrocardiogram and echocardiography tests, and adverse event reporting. Secondary endpoints include expression of dystrophin protein, motor function assessment, exon 44 skipping efficiency, plasma and urinary NS-089/NCNP-02 concentrations, and changes in blood creatine kinase levels.

**Discussion:** Exon-skipping therapy using ASOs shows promise in selected patients, and this first-in-human study is expected to provide critical information for subsequent clinical development of NS-089/NCNP-02.

## Background

Globally, the X-linked recessive disorder Duchenne muscular dystrophy (DMD) is reported to occur with a birth prevalence of 19.8 per 100,000 males [1]. DMD is the most common form of childhood-onset muscular dystrophy, caused by mutations in the *DMD* gene that result in absent or insufficient levels of the functional cytoskeletal protein dystrophin [2]. This underlying pathophysiology results in progressive muscular degeneration and damage and, ultimately, early mortality [3].

Owing to its chromosomal localization, DMD predominantly affects male children, while females are generally (although not exclusively) asymptomatic carriers [4]. Clinical symptoms include muscle weakness and degeneration in early childhood; the diagnosis of DMD was based on the results of genetic analysis by multiplex ligation-dependent probe amplification (MLPA) and direct sequencing of the *DMD* gene [3]. By the age of 12, individuals with DMD are typically non-ambulant. They require a wheelchair [2] and commonly develop additional orthopedic problems such as scoliosis [5]. This, in turn, can lead to chest cavity reduction and respiratory dysfunction [2]. The development of cardiomyopathy in young adulthood may cause additional breathing difficulties [6]. As a result, for most individuals with DMD, the average life expectancy is approximately 30 years [3].

Until now, many available treatments (e.g., surgery for spinal deformity, cardiac failure management, respiratory support, physiotherapy) addressed only the symptoms of DMD and were able to slow the course of the disease but could not halt the progressive muscle loss [7]. The only accepted disease-modifying treatment has been the use of corticosteroids, which can suppress muscle inflammation; however, the efficacy of corticosteroid treatment is limited and side effects can be considerable [8, 9].

Based on the clear need for improved disease-modifying treatments for DMD, several radical therapies have been studied in recent years, including gene therapy and stem cell transplantation [10]. Exon-skipping therapy, which induces an in-frame mutation in mature messenger RNA (mRNA) with sequence-specific antisense oligonucleotides (ASOs), is of particular interest to DMD researchers and clinicians [7, 11]. Exon skipping aims to mask specific exons in the *DMD* gene transcripts, thereby overcoming the out-of-frame mutation that underlies DMD and restoring the expression of a shorter but functional dystrophin protein [12–14]. In-frame deletions result in the production of truncated dystrophin, which translates into a less severely affected Becker muscular dystrophy (BMD). Several exon-skipping therapeutic options have now been approved for use in the United States (US), although their availability elsewhere differs. Eteplirsen (Exondys 51) is an exon 51-skipping ASO that was approved to treat patients with DMD by the Food and Drug Administration (FDA) in the US in 2016 [15]; however, it has not yet been approved by the European Medicines Agency for use in Europe. Golodirsen (Vyondys 53) is an exon 53-skipping ASO approved by the FDA in 2019 [16]. Viltolarsen (NS-065/NCNP-01; Viltepso) is another exon 53-skipping ASO, and this treatment was approved in 2020 by both the FDA [17] and the Japanese Pharmaceuticals and Medical Devices Agency [18].

Viltolarsen (NS-065/NCNP-01) was discovered and developed by the National Center of Neurology and Psychiatry (NCNP) in collaboration with Nippon Shinyaku Co., Ltd. In preclinical studies, this morpholino oligomer was found to promote skipping of exon 53 in a dose-dependent manner and restore dystrophin protein levels in patient-derived cells [19]. Based on the reported locations of mutations in the *DMD* gene, it was anticipated that NS-065/NCNP-01 could benefit the 6%–9.4% of patients with DMD amenable to exon 53 skipping [20], and early-stage clinical development began in 2016 [21]. Based on positive phase II data [22], the FDA granted conditional approval to viltolarsen in 2020, pending the ongoing phase III RACER53 trial [23].

Subsequently, the NCNP researchers and Nippon Shinyaku Co., Ltd. are collaborating to develop another novel morpholino oligomer, NS-089/NCNP-02, which induces exon 44 skipping to correct the open reading frame. This exon was selected because the neighboring exon 45 is the single exon most commonly deleted [24]; in theory, in patients with exon 45 mutations, skipping of exon 44 should restore the open reading frame to allow translation of a partially functional dystrophin protein and result in less severe disease [24]. It will be anticipated that NS-089/NCNP-02 could benefit the 7%–11% of patients with DMD amenable to exon 44 skipping [25]. To verify the exon-skipping efficiency of NS-089/NCNP-02 in cells derived from DMD patients, primary fibroblasts were obtained from patients with DMD amenable to exon 44 skipping. After patient fibroblasts were transfected with the human MYOD gene to induce differentiation into myotubes, the myotubes were treated with NS-089/NCNP-02 in the presence of Endo-Porter as a delivery agent, and the effects of exon 44 skipping were measured by RT-PCR 2 days after the start of treatment. We confirmed NS-089/NCNP-02 induced efficient exon 44 skipping in cells from a patient with a deletion of exons 45 or exons 45-54 (i.e., a complete inability to produce functional dystrophin) (NCNP and Nippon Shinyaku Co., Ltd., data on file). Based on these results, there is an expectation that NS-089/NCNP-02 could induce dystrophin expression and suppress disease progression in patients amenable to exon 44 skipping (under review).

For trials of radical therapies in DMD, there is still an ongoing debate about the best method for showing clinical benefit [7, 26]. In general, the usefulness of an exon-skipping drug should be based on demonstrating its exon-skipping efficiency and its effect on dystrophin restoration. Thus, we planned this phase I/II study to undertake an early-phase clinical evaluation of NS-089/NCNP-02 in a limited number of patients. One of the specific objectives of this study was to assess the potential utility of NS-089/NCNP-02 as a drug for the treatment of patients with DMD by evaluating its safety and pharmacokinetics (PK) (manuscript in preparation). Additionally, we aimed to identify markers predictive of therapeutic efficacy and determine the optimal dosing for future studies.

## Methods/design

### Study design and setting

This is an open-label, dose-escalation, two-center phase I/II trial. The study is registered at clinicaltrials.gov under the identifier NCT04129294 and with the University hospital Medical Information Network as UMIN000038505. The study has been conducted at the NCNP (Tokyo, Japan) since 2 December 2019 and at Kagoshima University Hospital (Kagoshima, Japan) since January 2021, and the study was completed in 31 May 2022. The study site will be responsible for compensation in the event of a health injury that occurs to a subject as a result of participating in the present study. Any protocol modifications or amendments will require approval from the Institutional Review Board.

The trial was designed in two parts. Part 1 is a stepwise dose-finding stage (4 weeks). Part 2 is a 24-week evaluation period based on the dosages determined during Part 1. Interim assessments by the safety monitoring committee are planned to decide whether to transition between dose levels in Part 1 and proceed from Part 1 to Part 2; the relevant criteria are described in **Table 1**.

**Table 1.**
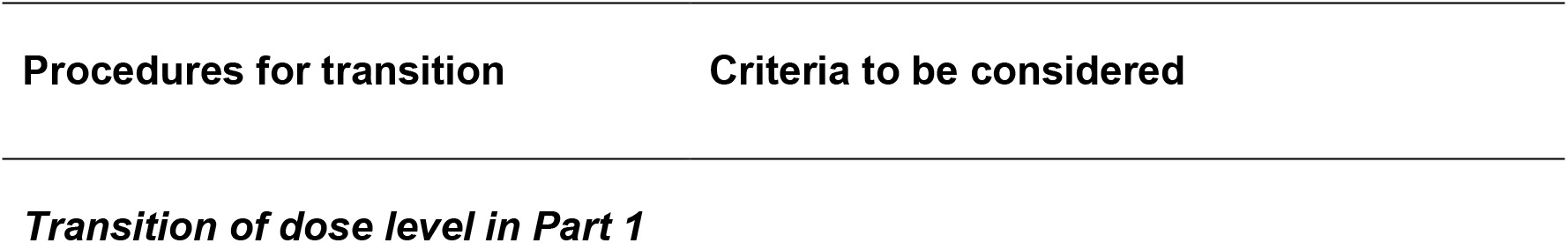
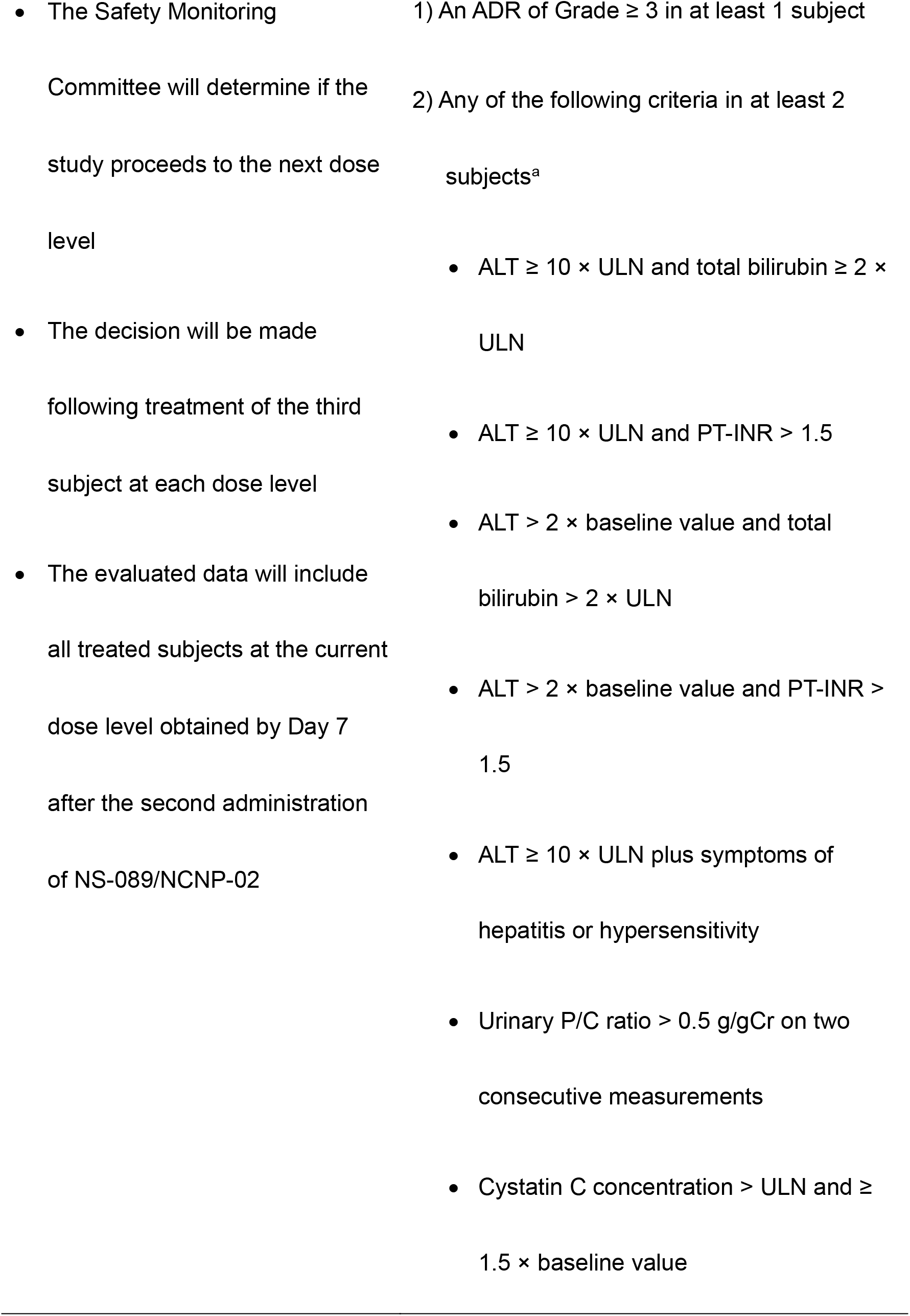
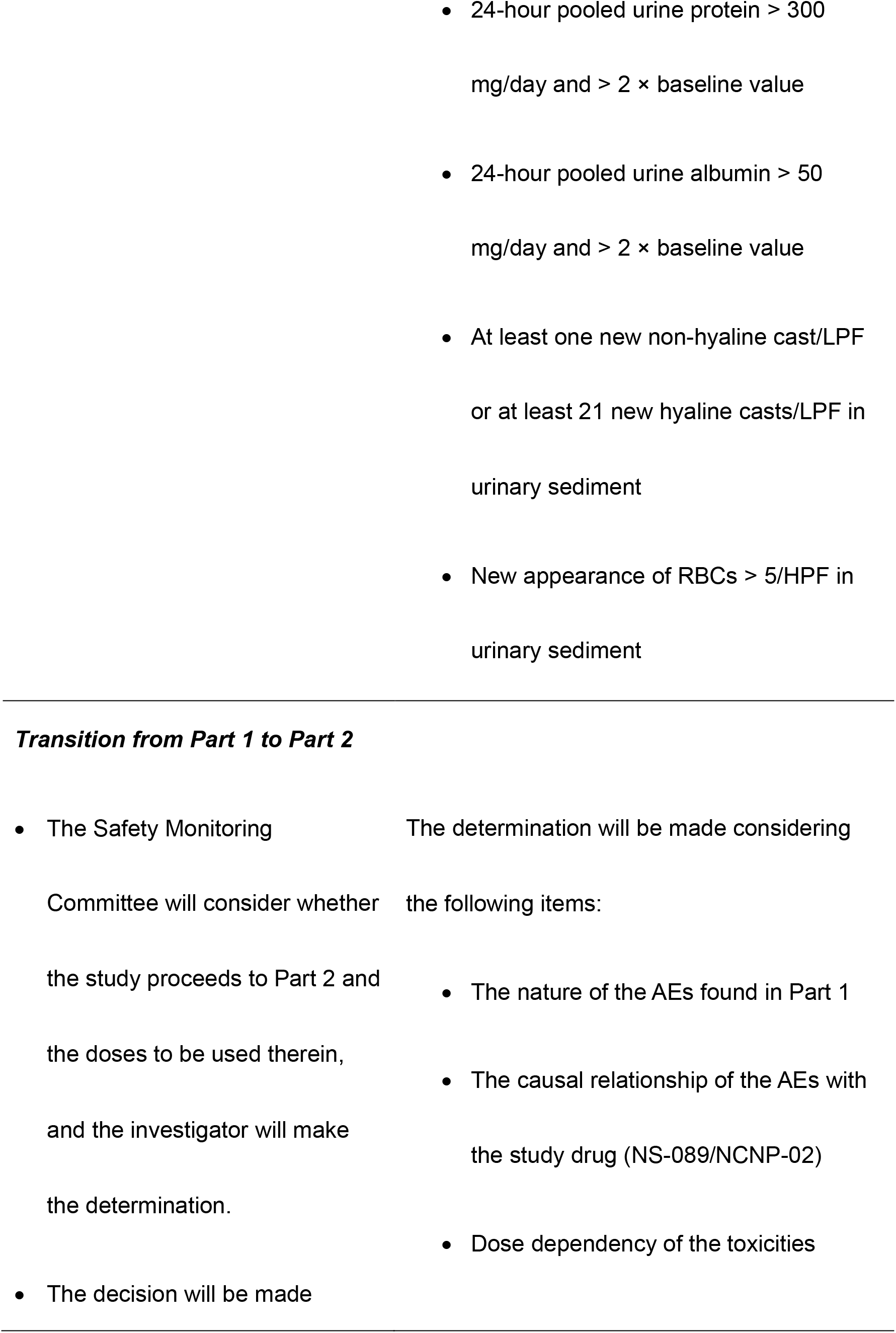
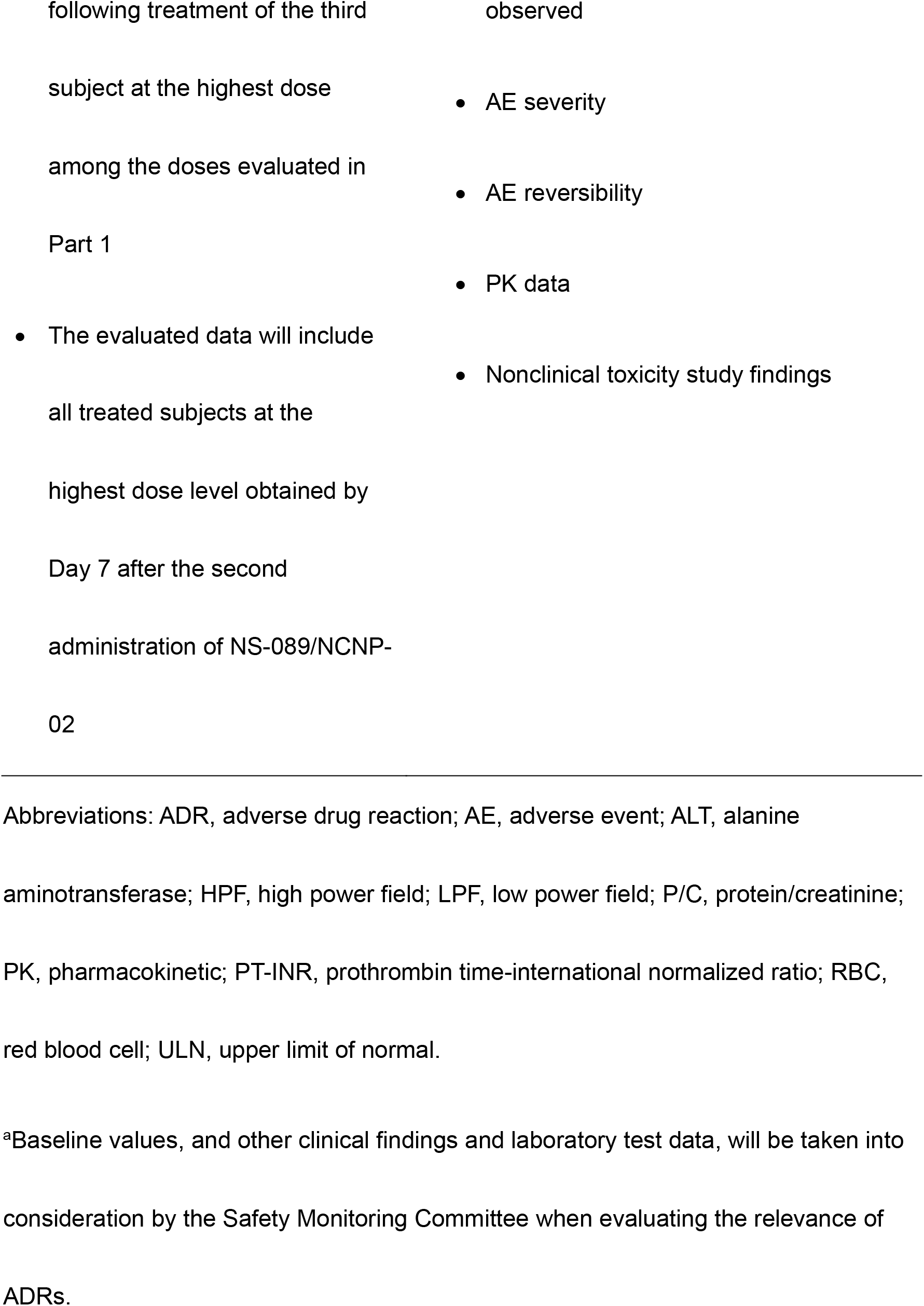
Criteria to determine whether to proceed to the next dose level and study part

### Study population

Patients enrolled in the study will be assigned subject identification codes to ensure subject confidentiality, and these codes will be used in case report forms and all documents related to the conduct of the study.

The study population will include ambulant patients (although non-ambulant patients may be enrolled depending on enrollment status) with DMD, presence of an out-of-frame deletion (confirmed by MLPA), and a dystrophin mutation amenable to exon 44 skipping (confirmed by direct sequencing). The key inclusion criteria are male patients who are ≥ 4 and < 17 years of age at the time of obtaining informed consent and who have a life expectancy of ≥ 1 year; presence of an out-of-frame deletion (confirmed by MLPA) that can be restored to an in-frame deletion by exon 44 skipping of the *DMD* gene; ability to undergo muscle biopsy (i.e., lacking severe atrophy of the biceps brachii muscle or tibialis anterior muscle); corrected QT interval (QTc) < 450 msec on standard 12-lead electrocardiogram (ECG) (based on Fridericia’s formula), or QTc < 480 msec for patients with bundle branch block; and systemic corticosteroid therapy (if any) started ≥ 6 months prior to enrollment with the dosage unchanged within 3 months of enrollment.

The study exclusion criteria include participation in any other clinical trial designed to restore the expression of dystrophin protein or related substances, or receipt of any other investigational drug within 3 months before the start of administration of NS-089/NCNP-02; impaired respiratory function to the extent that continuous use of a ventilator (excluding non-invasive positive pressure ventilation while sleeping) is necessary; a forced vital capacity of < 50% of the predicted value (based on the reference values for spirometry in Japanese children [27]), a left ventricular ejection fraction of < 40%, or fractional shortening of < 25% by ECHO; presence of any immunodeficiency, autoimmune disease, active or uncontrolled infection, cardiomyopathy, hepatic disease, renal disease, a positive test result for hepatitis B virus surface antigen, hepatitis C virus antibody, or human immunodeficiency virus antibody; any recent (within 3 months of enrollment) or planned surgery; a history of serious hypersensitivity reactions to pharmacologic drugs; and any patient unable to comply with the study procedures due to cognitive challenges or any patient whose safety might be compromised by study participation.

### Study treatment

During the study, NS-089/NCNP-02 is planned to be administered at four dose levels (1.62 mg/kg once weekly [QW], 10 mg/kg QW, 40 mg/kg QW, and 80 mg/kg QW). Dosing Level 1 (1.62 mg/kg QW) was selected as the first dose based on the results of animal toxicity studies (NCNP and Nippon Shinyaku Co., Ltd., data on file). Dosing Level 4 (80 mg/kg QW) was calculated based on a combination of data from the toxicity studies in animals (NCNP and Nippon Shinyaku Co., Ltd., data on file) and PK inferences from the related morpholino oligomer viltolarsen (NS-065/NCNP-01) administered at a dose of 80 mg/kg in humans [21]. The area under the plasma concentration-time curve from 0–24 hours (AUC_0-24_) when viltolarsen was administered at 80 mg/kg in humans did not exceed that of NS-089/NCNP-02 administered at the maximum tolerated dose in animals. It was, therefore, considered possible to administer NS-089/NCNP-02 at a dose of up to 80 mg/kg QW. NS-089/NCNP-02 will be administered intravenously over 1 hour (± 10 minutes).

### Study procedures

In Part 1 of the study, Cohort 1 (three patients) will initially receive NS-089/NCNP-02 at dosing Level 1 (1.62 mg/kg QW) for two weeks and at dosing Level 3 thereafter (40 mg/kg QW) for two weeks (**Figure 1**). Cohort 2 (three patients) will initially receive NS-089/NCNP-02 at dosing Level 2 (10 mg/kg QW) for two weeks and at dosing Level 4 thereafter (80 mg/kg QW) for two weeks. In Part 2 of the study, two different doses of NS-089/NCNP-02 (determined from the results of Part 1) will be intravenously administered QW for 24 weeks. Patients who have completed Part 1 are allowed to participate in Part 2. A muscle biopsies will be performed twice, at the beginning of Part 1 and at the end of Part 2.

**Figure 1.**
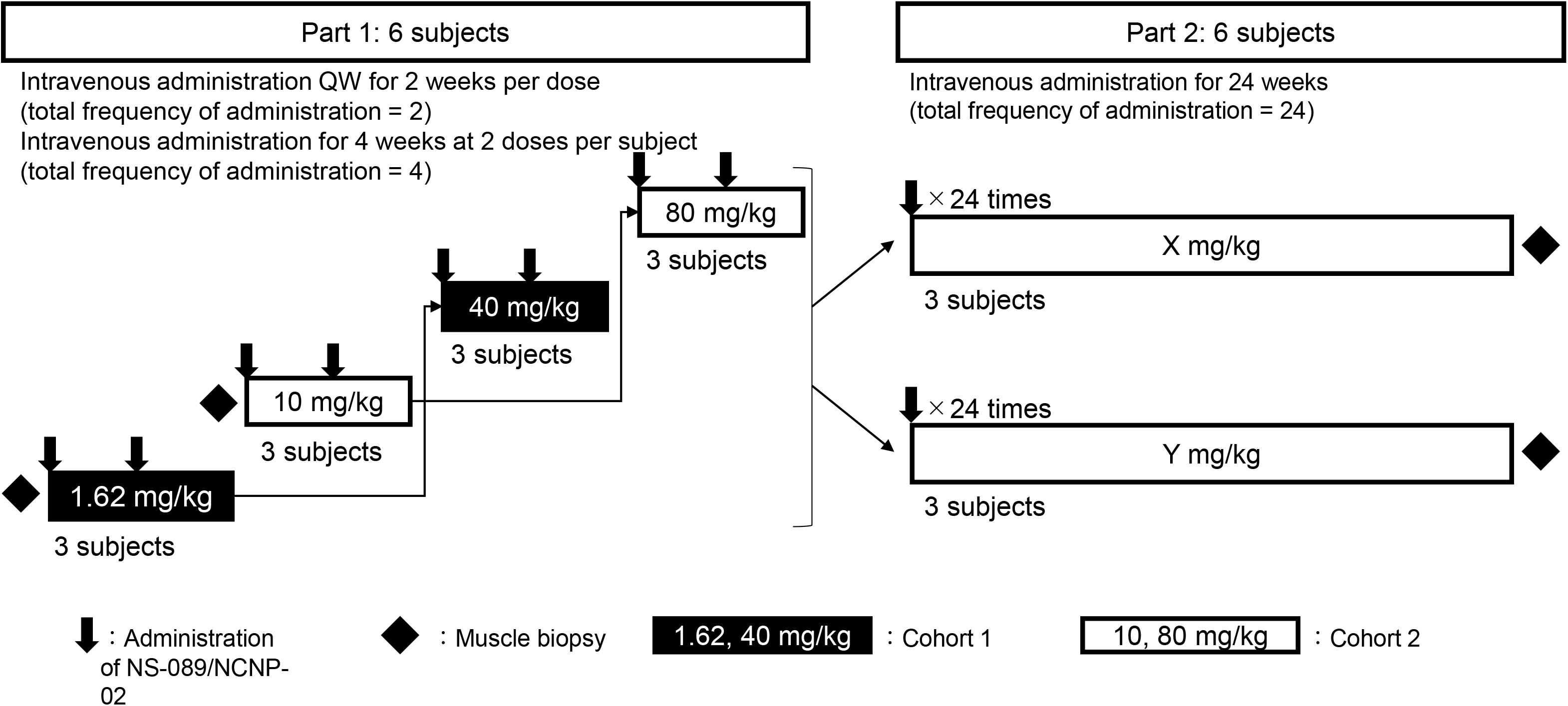
Study design and flow In Part 1, the dose of NS-089/NCNP-02 will be escalated in a stepwise manner from Level 1 (1.62 mg/kg QW) to Level 4 (80 mg/kg QW). A total of four doses will be intravenously administered to two cohorts consisting of three subjects each, and each cohort will receive two doses. In Part 2, two doses of NS-089/NCNP-02 (determined from the results of Part 1) will be intravenously administered once weekly for 24 weeks. Abbreviation: QW, once weekly.

If a patient discontinues, an additional subject will be added to evaluate three patients at each dose level. The second patient at each dose level in Part 1 will start receiving NS-089/NCNP-02 after the second administration of NS-089/NCNP-02 in the first patient. An interval of at least 4 weeks was mandated between dose levels in each cohort.

Physiotherapy or exercise therapy regimens must be continued without change between 90 days before starting the treatment and the end of the treatment period at Part 2; no new therapies can be initiated during the study period. Pre-existing concomitant drugs (including steroids) may be used between enrollment and the end of the treatment period at Part 2. Other investigational drugs, adenosine-triphosphate products, idebenone, and other coenzyme Q10 products will be prohibited. The schedule of observations and examinations is shown in **Figure 2**.

**Figure 2.**
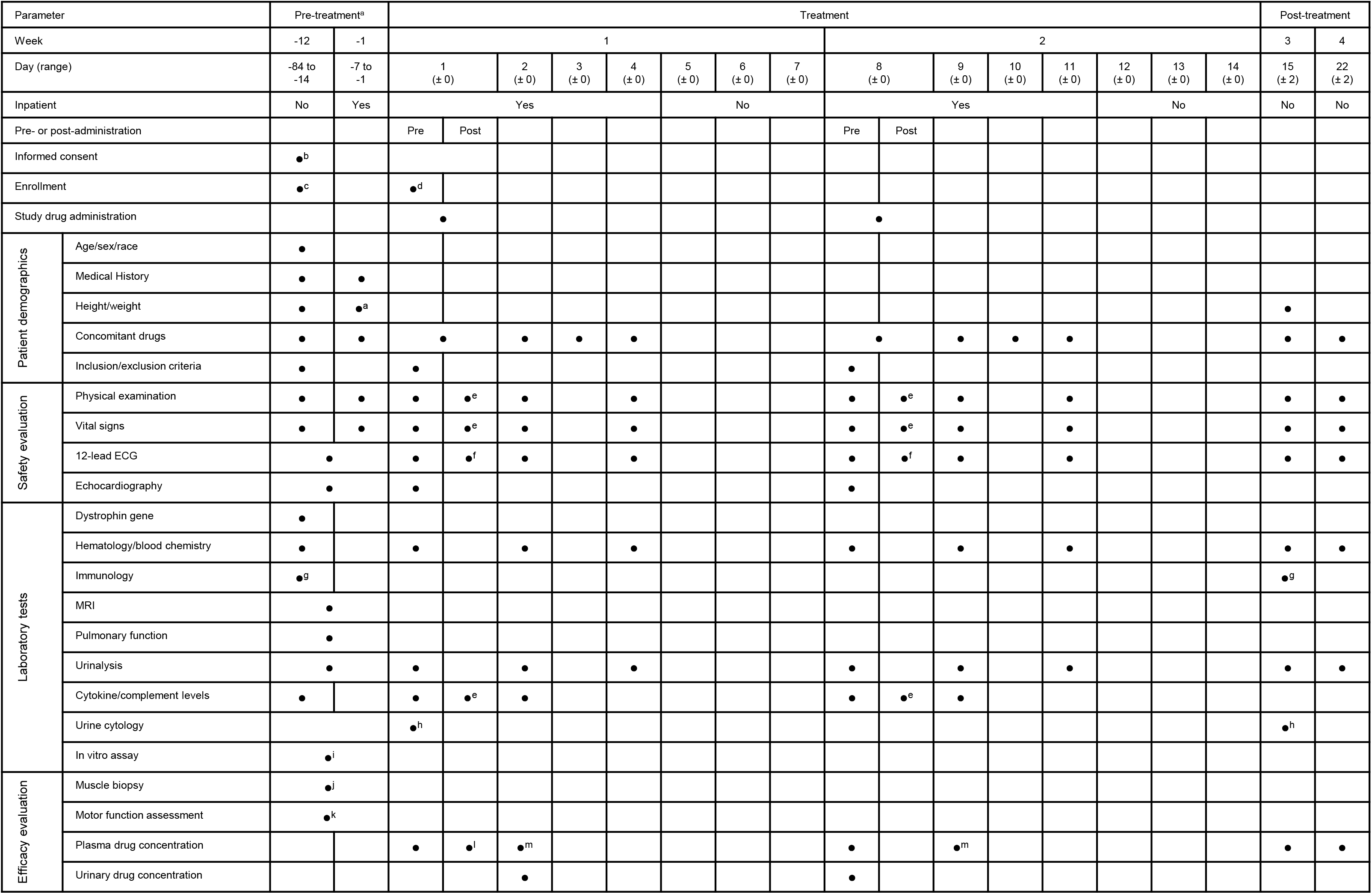
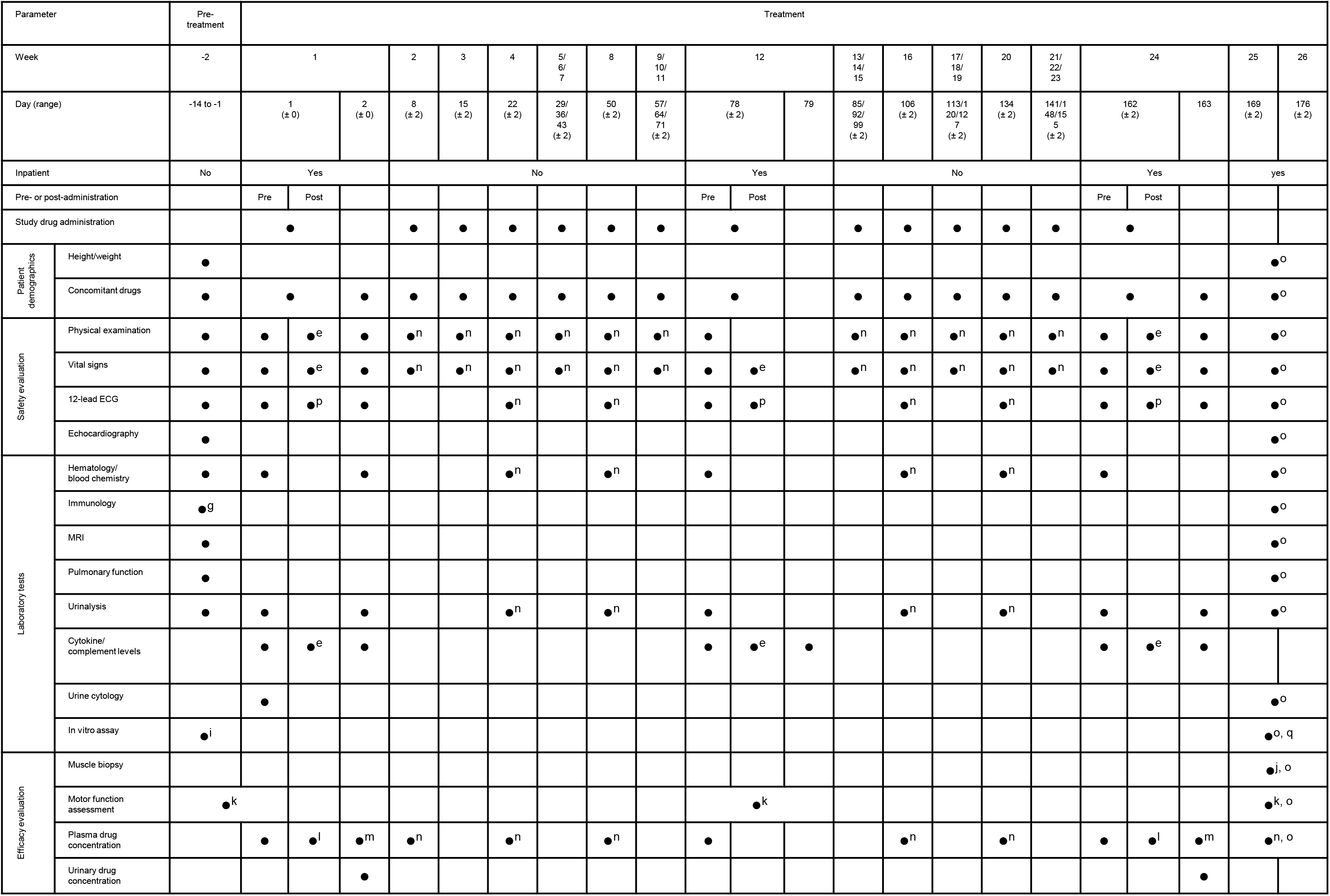
Study observations and examinations: implementation schedules. **a**: Part 1; **b**: Part 2 ^a^Patients who complete Level 1 or 2 and proceed to Level 3 or 4 will not have a pre-treatment observation period prior to the administration at Level 3 or 4, but will have their height/weight measured during the period between Days −7 and −1 before administration of NS-089/NCNP-02. ^b^Consent may be obtained before the day of interim enrollment. ^c^Interim enrollment. After obtaining informed consent, the investigator or sub-investigator will confirm that the patient is a candidate for exon 44 skipping treatment and enroll the patient. The results of exon 44 sequence analysis shall be confirmed before formal enrollment. ^d^Formal enrollment. Patients will be enrolled after confirming examination results obtained within 14 days before the start of administration of NS-089/NCNP-02. Formal enrollment on the day before the first administration may be accepted. ^e^To be performed at the end of administration and at 2 hours after the end of administration. ^f^To be performed at the following times: 30 minutes after the start of administration on Day 1; at the end of administration on Day 1; at 1, 2, 4, and 6 hours after the end of administration on Day 1; at the end of administration on Day 8; and at 1 hour after the end of administration on Day 8. ^g^For Part 1, the results of hepatitis B antigen, hepatitis C antibody, and human immunodeficiency virus antibody tests must be confirmed by the day of formal enrollment; such tests will not be performed post-treatment for Part 1 and pre-treatment for Part 2. ^h^To be performed at Levels 3 and 4. ^i^Muscle and skin samples will be taken simultaneously with muscle biopsy. Urine samples will be collected at Part2, if it could not be collected in a pre-treatment observation period at Part 1. ^j^To be performed after implementing motor function assessment and confirming the muscle tissue to be sampled by MRI. In addition, this is to be performed before the start of administration (pre-treatment for Part 1). Subjects will be admitted at the time of muscle biopsy. ^k^In Part 1, the motor function assessment will be performed between Days −28 and −1. In Parts 1 and 2, the 2-minute walk test will be conducted on a different day from other motor function parameters. ^l^To be performed 30 minutes after the start of administration, at the end of administration, and at 15 minutes, and 1, 2, 4, and 8 hours after the end of administration. ^m^To be performed 23 hours after the end of administration. ^n^To be performed before administration. °To be performed on Day169 or 176. ^p^To be performed at the end of administration and 1 hour after the end of administration. ^q^Muscle sample is collected at the same time as muscle biopsy. Abbreviations: ECG, electrocardiogram; MRI, magnetic resonance imaging.

### Study endpoints

The primary (safety) endpoints are the results of physical examinations, vital signs, 12-lead ECG, echocardiography tests, and adverse event (AE) reporting. Measures include *DMD* gene tests (MLPA analysis), exon 44 sequencing analysis (by polymerase chain reaction using genome DNA derived from peripheral venous blood), hematology, blood chemistry, urinalysis, cytokine/complement levels (**Table 2**), immunology tests, and pulmonary function tests. AEs and adverse drug reactions (ADRs) will be coded using the Medical Dictionary for Regulatory Activities version 23.1. Blood samples, muscle tissues, dermal fibroblasts, and urine-derived cells collected will be anonymized and stored at a genetic testing laboratory or NCNP with no time limit.

**Table 2.**
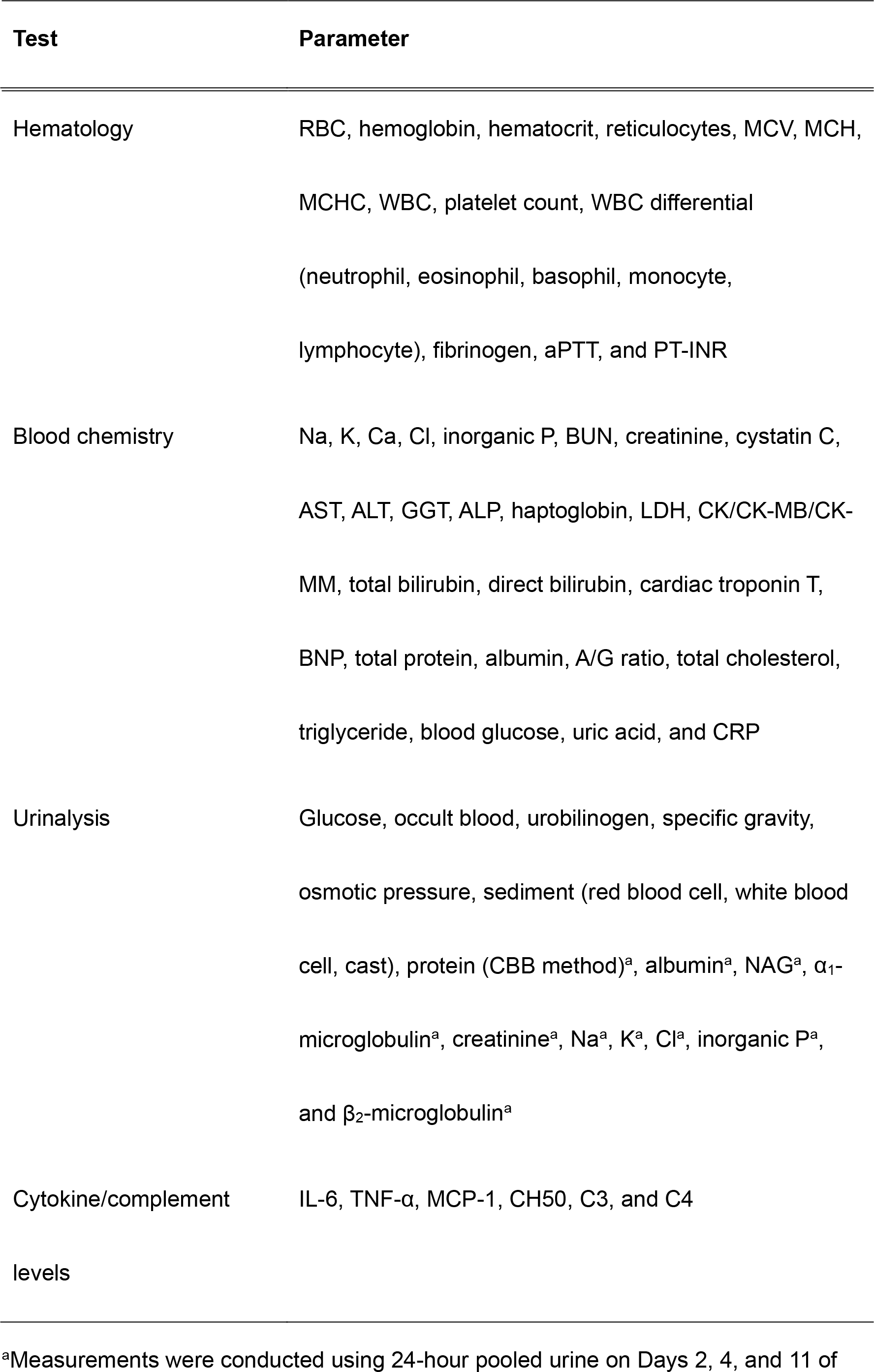
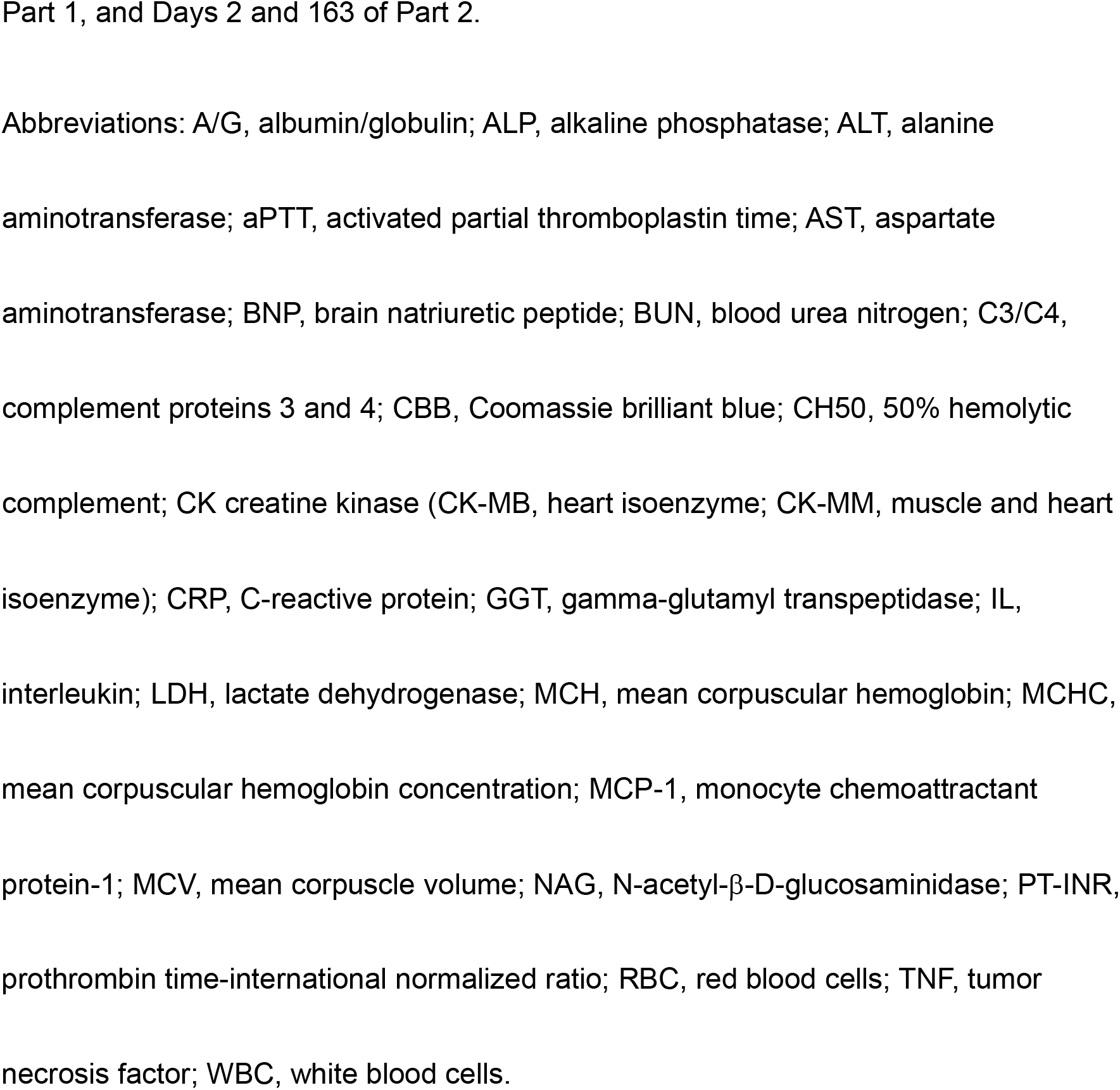
Laboratory test measures

| Test | Parameter |
| --- | --- |
| Hematology | RBC, hemoglobin, hematocrit, reticulocytes, MCV, MCH, MCHC, WBC, platelet count, WBC differential (neutrophil, eosinophil, basophil, monocyte, lymphocyte), fibrinogen, aPTT, and PT-INR |
| Blood chemistry | Na, K, Ca, Cl, inorganic P, BUN, creatinine, cystatin C, AST, ALT, GGT, ALP, haptoglobin, LDH, CK/CK-MB/CK-MM, total bilirubin, direct bilirubin, cardiac troponin T, BNP, total protein, albumin, A/G ratio, total cholesterol, triglyceride, blood glucose, uric acid, and CRP |
| Urinalysis | Glucose, occult blood, urobilinogen, specific gravity, osmotic pressure, sediment (red blood cell, white blood cell, cast), protein (CBB method)<sup>a</sup>, albumin<sup>a</sup>, NAG<sup>a</sup>, $\alpha_1$-microglobulin<sup>a</sup>, creatinine<sup>a</sup>, Na<sup>a</sup>, K<sup>a</sup>, Cl<sup>a</sup>, inorganic P<sup>a</sup>, and $\beta_2$-microglobulin<sup>a</sup> |
| Cytokine/complement levels | IL-6, TNF-α, MCP-1, CH50, C3, and C4 |

<sup>a</sup>Measurements were conducted using 24-hour pooled urine on Days 2, 4, and 11 of
Part 1, and Days 2 and 163 of Part 2.
Abbreviations: A/G, albumin/globulin; ALP, alkaline phosphatase; ALT, alanine
aminotransferase; aPTT, activated partial thromboplastin time; AST, aspartate
aminotransferase; BNP, brain natriuretic peptide; BUN, blood urea nitrogen; C3/C4,
complement proteins 3 and 4; CBB, Coomassie brilliant blue; CH50, 50% hemolytic
complement; CK creatine kinase (CK-MB, heart isoenzyme; CK-MM, muscle and heart
isoenzyme); CRP, C-reactive protein; GGT, gamma-glutamyl transpeptidase; IL,
interleukin; LDH, lactate dehydrogenase; MCH, mean corpuscular hemoglobin; MCHC,

Planned secondary endpoints include dystrophin protein expression (by western blot analysis), motor function assessment, exon 44 skipping efficiency, plasma and urinary NS-089/NCNP-02 concentrations, and changes in blood creatine kinase levels. Motor function assessments include the North Star Ambulatory Assessment score [28]; the time to stand from supine test [29]; the 10-meter walk/run test [30]; the 6-minute walk test [31]; the 2-minute walk test [32]; the timed “up & go” test [33]; quantitative muscle strength assessment (knee flexion/extension, elbow flexion/extension, and shoulder abduction); and grip strength, pinch strength, and upper limb performance assessed using the Performance of the Upper Limb module for DMD [34].

Additional study examinations included skeletal muscle magnetic resonance imaging (to determine the amount of muscle that can be biopsied); in vitro assays using MYOD-transduced urine-derived cells (MYOD-UDCs), dermal MYOD-fibroblasts, and myoblasts derived from the six enrolled subjects in the trial, evaluated by dystrophin RT-PCR and western blot analysis; and plasma and urinary PK parameters. The timings of the blood and urine collections are shown in **Additional file 1**. NS-089/NCNP-02 concentration and PK parameters will be measured by high-performance liquid chromatography with tandem mass spectrometry.

### Statistical analysis

The sample size in each study part was set at six patients. This sample size was determined based on the feasibility of enrollment and motor function assessment at a single center. No statistical significance was considered for the determination of sample size. Data will be assessed in three analysis sets (safety, efficacy, and PK). The safety analysis set will include all patients treated at least once with NS-089/NCNP-02. The efficacy and PK analysis sets will include all patients enrolled, excluding those who have a serious eligibility violation and those who lack the applicable data.

Patient demographic data will be tabulated using summary statistics. The primary analysis (safety evaluation) will include tabulation of AEs and ADRs by number and frequency and graphical representation of changes from baseline in vital signs, mean laboratory test values, ECG, echocardiography, and pulmonary function parameters. The efficacy and PK analyses will include an itemization of motor function testing and calculation of NS-089/NCNP-02 exposure (including the maximum plasma concentration [C_max_] and AUC_0-24_), half-life, systemic clearance, and distribution volume. Analyses will also include tabulation of administration of NS-089/NCNP-02 (number of administrations and exposure level by dose in each of the study parts). Analyses will be conducted using SAS software version 9.4 (SAS Institute Inc., Cary, NC, USA) and Microsoft Excel 2016/2019.

### Data control and dissemination

All data and processes will be audited by independent monitors. The study data will be published in peer-reviewed journals, using anonymized information to preserve patient confidentiality.

## Discussion

This is an exploratory phase I/II study of NS-089/NCNP-02. The protocol is reported according to the Standard Protocol Items: Recommendation for Interventional Trials (SPIRIT) 2013 Checklist (**Additional File 2**).

DMD is a disease with a significant impact on life expectancy, and although radical therapies are being investigated, these generally involve complex procedures and other challenges. Exon-skipping therapy using ASOs is a treatment that shows promise in selected patients. This first-in-human study is expected to provide critical information for subsequent clinical development of NS-089/NCNP-02, with the ultimate aim of expanding the treatment armamentarium for affected patients.

In this study, the in vitro assay, which includes the differentiation of urine-derived cells into myocytes, is based on the authors’ recently published technique to create a novel DMD muscle cell model [35]. This method could be used to screen different ASOs prior to commencing clinical trials [20, 36], potentially streamlining the therapeutic development process and reducing the time to availability of new treatment strategies. Recently, we have reported an efficient cellular skeletal muscle modeling in DMD using MYOD-UDCs obtained from DMD patients [35]. UDCs might be an ideal cellular model for neuromuscular diseases because they have the high proliferative ability and can be collected non-invasively. However, only a few studies are conducted on MYOD-UDCs, it is desirable to use them in parallel with the well-characterized MYOD-fibroblasts model [36]. Very recently, we, for the first time, evaluated the usefulness of MYOD-UDCs from study subjects to evaluate the efficacy of NS-089/NCNP-02 in the DMD clinical trial. We confirmed the drug’s dose-dependent exon 44 skipping and dystrophin protein recovery in MYOD-UDCs obtained from all the subjects (manuscript in preparation).

The modelling of truncated dystrophin protein lacking exons 44-45 or 45-46 predicts that both should result in comparable stable hybrid rod structure, suggesting that, in patients with exon 45 mutations, skipping of exon 44 or exon 46 should be excellent therapeutic strategies [24]. However, it is already known that the resulting protein is functional enough to maintain a mild BMD phenotype, which generally has mutations in the *DMD* gene that maintain the open reading frame, allowing the production of dystrophin proteins and thus are partially functional [38]. A documented example of this is the spontaneous skipping of exon 44 that occurs when the exons flanking it are deleted [39, 40]. Importantly, deletions amenable to exon 44 skipping are usually associated with more dystrophin-revertant fibers and milder DMD phenotypes [41–43]. As the stability of the dystrophin transcript is a key indicator of dystrophin expression, additional clinical exploration is warranted to discern whether transcript instability in DMD compared with BMD could be a potential biomarker of response to ASOs [38].

Potential study limitations include the small sample size and open-label design, limiting the conclusions that can be drawn. Then, significant challenges about ASOs are the difficulty in selecting an optimal sequence for exon-skipping drug, identifying biomarkers of treatment effect, large-scale pharmaceutical synthesis, and pharmaceutical marketing. Other limitations are the requirement for more sensitive outcome measures to quantify disease severity and our current incomplete understanding of the long-term natural history of DMD in ambulant patients with mutations amenable to exon 44 skipping.

The duration of the Part2 is short for identify the predictive efficacy, although the progression of DMD in exon 44-skipped patients is relatively slow. Then, the long-term extension study for 72 weeks is ongoing by our collaborator Nippon Shinyaku (NCT05135663).

In summary, in this phase I/II study, we plan to assess the safety and PK of the novel morpholino oligomer, NS-089/NCNP-02, which induces exon 44 skipping in the *DMD* gene. These data will allow us to determine the optimal dosage for future clinical development and provide the first indication of therapeutic efficacy. As such, the safety, efficacy, and dosing analyses of this study are critical precursors to undertaking later phase studies of NS-089/NCNP-02.

### Trial status

The study was completed on 31 May 2022. The first patient was included on 2 December 2019. Enrollment is now finished, with the final patient’s enrollment on 24 March 2020. The current article is based on protocol version 12.0, prepared on 11 November 2021.

## Supplementary Information

**File name:** Additional file 1

File format: .doc

**Title:** Collection of blood and urine for measurement of NS-089/NCNP-02 concentration and PK parameters.

**Description:** This table shows the timings of the blood and urine collections.

**File name:** Additional file 2

File format: .doc

**Title:** SPIRIT checklist.

## Abbreviations

ADR: adverse drug reaction
AE: adverse event
ASO: antisense oligonucleotide
AUC0-24: area under the plasma concentration-time curve from 0–24 hours
BMD: Becker muscular dystrophy
Cmax: maximum plasma concentration
DMD: Duchenne muscular dystrophy
ECG: electrocardiogram
ECHO: echocardiogram
FDA: Food and Drug Administration
MLPA: multiplex ligation-dependent probe amplification
mRNA: messenger ribonucleic acid
NCNP: National Center of Neurology and Psychiatry
PK: pharmacokinetics
QTc: corrected QT interval
QW: once weekly
US: United States

## Data Availability

The study data will be published in peer-reviewed journals, using anonymized information to preserve patient confidentiality.

## Acknowledgments

The authors would like to thank the patients and their families for participating in this study. The authors also thank Sally-Anne Mitchell, PhD, of Edanz Pharma for providing medical writing services funded by Nippon Shinyaku Co., Ltd., Kyoto, Japan.

## Disclosure

### Data Availability Statement

The datasets are available from the corresponding author upon reasonable request.

### Approval of the research protocol by an Institutional Reviewer Board

This study is being conducted in compliance with the ethical principles that have their origin in the Declaration of Helsinki and all applicable Japanese local and national regulatory laws. The protocol and related documentation were approved by the Institutional Review Board of the NCNP (approval number II-012).

### Informed Consent

Legal representatives for each patient provided written informed consent prior to study participation, and patients provided voluntary assent where possible.

### Registry and the Registration No. of the study/trial

This study was registered at clinicaltrials.gov (NCT04129294) and with the University Hospital Medical Information Network (UMIN000038505).

### Authors’ contributions

TI, HK, YAs, HN, and YAo conceived and designed the study and were involved in protocol development. TI, YAs, and HN coordinated the regulatory aspects. ET, YSM, AI, CY and SM were responsible for data acquisition. TI, HK, and EH were responsible for data analysis and interpretation. TI, HK, and YAo wrote the first draft of the manuscript. All other authors critically reviewed the manuscript for intellectual content. All authors read and approved the final version of the manuscript.

